# Genetic variation within the human papillomavirus type 16 genome is associated with oropharyngeal cancer prognosis

**DOI:** 10.1101/2021.11.11.21266230

**Authors:** Krystle A. Lang Kuhs, Daniel Faden, Li Chen, Derek K. Smith, Maisa Pinheiro, C. Burton Wood, Seth Davis, Meredith Yeager, Joseph F. Boland, Michael Cullen, Mia Steinberg, Sara Bass, Xiaowei Wang, Ping Liu, Mitra Mehrad, Thomas Tucker, James S. Lewis, Robert L. Ferris, Lisa Mirabello

**Author notes:** The last two authors contributed equally to this work. **Corresponding author:** Krystle A. Lang Kuhs, PhD, MPH, Associate Professor, Department of Epidemiology, College of Public Health, University of Kentucky, 760 Press Ave, Room, Lexington, KY 40508, USA.

## Abstract

**Purpose:** A significant barrier to adoption of de-escalated treatment protocols for human papillomavirus-driven oropharyngeal cancer (HPV-OPC) is that few predictors of poor prognosis exist. We conducted the first large whole-genome sequencing (WGS) study to characterize the genetic variation of the HPV16 genome and to evaluate its association with HPV-OPC patient survival.

**Patients and Methods:** 460 OPCs from 2 large US medical centers (1980-2017) underwent HPV16 WGS. Site-specific variable positions (SNPs) across the HPV16 genome were identified. Cox proportional hazards models estimated hazard ratios (HRs) and 95% confidence intervals (CIs) for overall survival by HPV16 SNPs. Harrell C-index and time-dependent positive predictive value (PPV) curves and areas under the PPV curves were used to evaluate the predictive accuracy of HPV16 SNPs for overall survival.

**Results:** 384 OPCs (83.48%) passed quality control filters with sufficient depth and coverage of HPV16 genome sequencing to be analyzed. 284 HPV16 SNPs with a minor allele frequency >1% were identified. Eight HPV16 SNPs were significantly associated with worse survival after false discovery rate (FDR) correction (individual prevalence:1.0%-5.5%; combined prevalence: 15.10%); E1 gene position 1053 (HR for overall survival [HR_os_]:3.75,95%CI:1.77-7.95;*P*_fdr_=0.0099); L2 gene positions 4410 (HR_os_:5.32,95%CI:1.91-14.81;*P*_fdr_=0.0120), 4539 (HR_os_:6.54,95%CI:2.03-21.08;*P*_fdr_=0.0117); 5050 (HR_os_:6.53,95%CI:2.34-18.24;*P*_fdr_=0.0030) and 5254 (HR_os_:7.76,95%CI:2.41-24.98;*P*_fdr_=0.0030); and L1 gene positions 5962 (HR_os_:4.40,95%CI:1.88-10.31;*P*_fdr_=0.0110) and 6025 (HR_os_:5.71,95%CI:2.43-13.41;*P*_fdr_=0.0008) and position 7173 within the upstream regulatory region (HR_os_:9.90,95%CI:3.05-32.12;*P*_fdr_=0.0007). Median survival time for patients with ≥1 high-risk HPV16 SNPs was 3.96 years compared to 18.67 years for patients without a high-risk SNP; log-rank test *P*<0.001. HPV16 SNPs significantly improved the predictive accuracy for overall survival above traditional factors (age, smoking, stage, treatment): increase in C-index was 0.069 (95% CI: 0.019-0.119, *P* <0.001); increase in area under the PPV curve for predicting 5-year survival was 0.068 (95%CI: 0.015-0.111, *P* =0.008).

**Conclusions:** HPV16 genetic variation is associated with HPV-OPC prognosis and can improve prognostic accuracy.

## INTRODUCTION

The incidence of oropharyngeal cancer (OPC) is rapidly increasing within the US due to an increase in the subset caused by human papillomavirus infection (HPV-OPC).^1-8^ HPV is now responsible for approximately 75% of all OPCs within the US.^7, 9^ Approximately 90% of HPV-OPC cases are due to just one HPV type – HPV16.^10, 11^

HPV positivity is the single most important prognostic factor for survival; 3-year overall survival is 82% among HPV-OPC patients compared to 57% for HPV-negative OPC patients.^12^ Despite high survival rates, HPV-OPC treatment is associated with severe morbidity including speech problems, difficulties in swallowing/eating, and chronic pain.^13^ Considerable interest is focused on de-escalation of treatment to reduce treatment-associated morbidity. However, not all patients with HPV-OPC have favorable outcomes; up to 25% recur within 3 years, and following recurrence, 2-year survival is only 55%.^14, 15^ The most significant barrier for the adoption of de-escalated treatment protocols is the inability to identify patients at high risk for recurrence and death. Currently, there are no clinically utilized markers for HPV-OPC prognosis.

A genetic characterization of 279 head and neck cancers by The Cancer Genome Atlas found that HPV-driven tumors had significantly fewer genetic alterations than HPV-negative tumors suggesting that the HPV genome may be a potential driver of tumorigenesis and; thus, potentially an important factor for treatment response and/or recurrence.^16^ Yet, no study to date has evaluated whether genetic variation within the HPV genome explains why a subset of patients have poor outcomes. However, recent advances in HPV whole genome sequencing (WGS) technology is allowing for the HPV16 genome to be interrogated at a level of detail impossible just a few years ago. In a large study of over 5,500 HPV16 whole-genomes from cervical specimens, our group found that extreme genetic conservation of the viral E7 oncogene and HPV16 SNPs located within the upstream regulatory region (URR) were strongly associated with risk of developing cervical precancer and cancer.^17^

We recently expanded upon our prior work by using next-generation HPV16 whole genome sequencing to conduct the first large study to characterize the genetic variation of the HPV16 genome within HPV-OPC.^18-20^ In total, we evaluated 460 OPC tumor specimens from 2 large US-based academic medical centers. Here, we present results pertaining to HPV-OPC patient prognosis. To our knowledge, this is the first study, at any anatomic site, to evaluate the predictive accuracy of HPV16 genomic variation and patient survival.

## MATERIALS AND METHODS

### Participants and Study Design

#### Vanderbilt University Medical Center (VUMC)

Incident, previously untreated cases of OPC were identified though the Vanderbilt Research Derivative (RD), an IRB-approved, identified, searchable database of more than 3.5 million electronic health records (EHRs) from patients seen at VUMC.^21^ The RD contains clinical data collected as part of routine patient care but reorganized to be easily searchable and usable for research purposes. The RD also links with the Vanderbilt Cancer Registry (VCR), which collects detailed clinical information from all reportable neoplasms diagnosed and/or treated at VUMC. Patients diagnosed between June 1, 2000 and July 9,2018 were identified using the following International Classification of Diseases for Oncology, 3^rd^ Edition (ICD-O-3) codes: C01.9, C02.4, C05.1, C05.2, C05.8, C09.0, C09.1, C09.8, C09.9, C10.0, C10.2, C10.3, C10.8, C10.9, C14.0 and C14.2. Clinical information prior to the year 2000 was not reliably captured within the VUMC EHR system; thus, cases diagnosed prior to 2000 were excluded. Each patient’s EHR was manually reviewed. Patients for whom an OPC diagnosis could not be confirmed, those with a prior history of cancer (other than non-melanoma skin cancer), patients who were immunocompromised, and those without a tumor specimen collected prior to treatment were excluded. For patients that met inclusion criteria, all surgical pathology specimen slides and pathology files created as part of the original diagnosis were reviewed by a trained head and neck pathologist (JSL). Tumor specimens of insufficient size for DNA extraction, those with original histologic diagnoses inconsistent with the findings of the pathology re-review, or with tumor blocks missing and/or exhausted were excluded. In total, 290 specimens were available for DNA isolation, of which, 280 were sequenced at NCI (Supplemental Figure 1). All cases, regardless of p16 test results, were sent to the NCI for sequencing. All studies were reviewed and approved by the Vanderbilt IRB.

#### University of Pittsburgh Medical Center (UPMC)

Incident, previously untreated, cases of OPC were identified using an IRB-approved protocol (UPCI 99-069) that collects biospecimens at the University of Pittsburgh Medical Center; all patients provided written informed consent. Since the establishment of the biobank, a total of 1,861 patients with OPC (ICD10 codes: C01.0, C02.4, C05.1, C05.2, C05.8, C09.0, C09.1, C09.8, C09.9, C10.0, C10.2, C10.3, C10.8, C10.9, C14.0) were treated at UPMC; 1225 (66%) were enrolled as part of the tissue banking study, of which, 180 (15%) were HPV-positive and had tumor DNA specimens available and were included in this study.

### Clinical Data Abstraction

Demographic and clinical information including treatment course, treatment responses, outcome (death) were obtained from each institutions’ cancer registry. An additional manual review of the EHRs was conducted to obtain data elements not routinely captured and/or frequently updated by the registries including p16 status, tobacco use, recurrence and last known follow up. For VUMC, study data were collected and managed using REDCap (Research Electronic Data Capture) electronic data capture tools hosted at Vanderbilt University.^22^ For UPMC, data were housed within the UPMC Head and Neck Biorepository.

### Specimen Collection, Processing and DNA isolation

Formalin fixed paraffin embedded (FFPE) specimens were retrieved from surgical pathology archives. Tumor regions were marked on the H&E slides by a head and neck pathologist for macro-dissection and DNA isolation was performed using a QiAamp DNA FFPE Tissue Kit (Qiagen) in accordance with the manufacturer’s instructions.

### p16 Immunohistochemistry

All UPMC specimens and a subset of VUMC specimens had prior p16 testing as part of routine clinical care. p16 immunohistochemistry was performed on all VUMC specimens without prior clinical testing using the E6H4 antibody (Ventana Medical Systems, Inc.). Slides were read by a head and neck pathologist (JSL) and interpreted as positive or negative according to CAP recommendations (positive = nuclear and cytoplasmic positivity in >70% of tumor cells of at least moderate to strong intensity).^23^

### HPV Genome Sequencing

Extracted tumor DNA was sent to the National Cancer Institute’s Cancer Genomics Research Laboratory (Rockville, Maryland) for sequencing. A custom Thermo Fisher Ion Torrent AmpliSeq HPV16 panel approach was used to amplify the HPV16 genome as previously described.^24^ This next-generation sequencing (NGS) assay uses Thermo Fisher Life Sciences’ Ion Torrent Proton in combination with a custom HPV16 Ion Ampliseq panel of 48 multiplexed primers designed to cover the entire viral genome for all HPV16 variant lineages. SNP calls were made using the Torrent Variant Caller v.5.0.3, and nucleotide variants were annotated with HPV gene/region using snpEff v.3.6c.^25^ Pipeline settings and parameters can be found at https://github.com/cgrlab/cgrHPV16. HPV16 lineage (A, B, C, D) and sublineage assignment (A1-4, B1-4, C1-4, D1-4) was based on the maximum likelihood (ML) tree topology using RAxML MPI v7.2.8.27^26^ and 16 HPV16 sublineage reference sequences. In the event that multiple HPV16 variants were detected, the predominant variant was assigned based on presence in at least 60% of the sequence reads. Seventy-two specimens were excluded due to poor read depth, incomplete coverage across the genome, or inability to assign lineage. This method was previously validated and was found to have 99.9% concordance with the “gold standard” Sanger sequencing.^24^

### Statistical Analyses

Patient characteristics were evaluated overall and by study site. The Cox proportional hazards models were used to evaluate the association between each individual HPV16 SNP (N=284 SNPs with a minor allele frequency >1%) and overall survival. To assess risk of death from any cause, the only censoring event was at time of last known follow-up; years since cancer diagnosis was used as the time variable. Multiple comparisons adjustment was performed to control the false discovery rate (FDR). Benjamini-Hochberg false discovery rates were calculated using the FDRestimation package in R. FDR-corrected *P*s less than 0.05 (2-tailed) were considered statistically significant. We further used the Harrell C-index^27^ and the time-dependent predictive curves^28, 29^ to evaluate the added predictive accuracy of the identified HPV16 SNPs for overall survival above traditional factors (age, smoking, stage, treatment). The Harrell C-index was used to evaluate the overall predictive accuracy for survival, and the time-dependent predictive curves were used to evaluate the predictive accuracy for survival at specific years. The time-dependent predictive curves extends the positive predictive value (PPV) and negative predictive value (NPV) for binary outcomes to time-to-event outcomes and characterize event/event-free probabilities at specific years for high/low risk subjects. Since we were interested in selecting patients at high risk, we used PPV curves in our analysis. We evaluated the added predictive accuracy of the identified HPV16 SNPs by comparing C-index and the 3-, 5-, and 8-year PPV curves between the Cox regression model with the identified HPV16 SNPs and traditional factors and the Cox model with traditional factors alone. The procedure to obtain the 3-, 5-, and 8-year PPV curves is as follows. First, under each Cox regression model, a risk score formula was constructed and a risk score was calculated for each subject. Second, for a given cut-off value of the risk score, the population was divided into high-risk (risk core ≥cut-off) and low-risk (risk core <cut-off) groups; the proportion of population in that high-risk group was calculated and the corresponding 3-, 5-, and 8-year death probabilities of that high-risk group were obtained by the Kaplan-Meier method. Finally, the 3-, 5-, and 8-year PPV curves were obtained by plotting the 3-, 5-, and 8-year death probabilities of the high-risk group against the proportion of population in that high-risk group, respectively. A higher PPV curve indicates a more predictive model. The difference in area under the PPV curve (ΔAUC) was calculated as a summary measure to compare the PPV curves of two models at a given year, which measures the averaged difference in probability of death at that given year between the high-risk groups selected by the two models. For both ΔAUC and increase in C-index, the 95% confidence intervals were calculated using the bootstrap method as in our previous publication.^29^ A 95% confidence interval with the lower limit greater than 0 indicates a statistically significant increase in predictive accuracy. Analyses were performed by STATA IC version 15 and R version 3.6.0.

## RESULTS

### Participant Characteristics

Of the 460 OPC tumors assessed, 384 (83.48%) passed our quality control filters with sufficient depth and coverage of HPV16 genome sequencing to be included in the analyses; 217 from VUMC and 167 from UPMC. Demographic and clinical characteristics were similar between study sites with the exception of year of diagnosis, smoking history, stage and treatment (Table 1). Overall, median age at diagnosis was 57 (interquartile range [IQR]: 51 to 64). The majority were male (89.32%), Caucasian (97.66%), reported a history of tobacco use (65.36%) and presented with stage I disease (68.23%; according to the 8^th^ edition of the American Joint Committee on Cancer [AJCC] guidelines). The vast majority of HPV16 lineages detected were A (90.36%). A1 was the most common HPV16 sublineage (55.99%; N=215) followed by A2 (27.86%; N=107; Table 1).

**Table 1:** Patient characteristics overall and by study

| Characteristics | Overall<br>N=384 | VUMC<br>N=217 | UPMC<br>N=167 | P-Value |
| --- | --- | --- | --- | --- |
|  | N (%) | N (%) | N (%) |  |
| Age at Diagnosis (years) |  |  |  |  |
| Median (IQR) | 57 (51-64) | 57 (50-65) | 57 (51-63) | 0.968 |
| Year of Cancer Diagnosis |  |  |  |  |
| 1980-1990 | 5 (1.30) | 0 (0.0) | 5 (2.99) |  |
| 1991-2000 | 17 (4.43) | 0 (0.0) | 17 (10.18) |  |
| 2001-2010 | 95 (24.74) | 90 (41.47) | 5 (2.99) |  |
| 2011-2018 | 267 (69.53) | 127 (58.53) | 140 (83.83) | <0.001 |
| Gender |  |  |  |  |
| Male | 343 (89.32) | 196 (90.32) | 147 (88.02) |  |
| Female | 41 (10.68) | 21 (9.68) | 20 (11.98) | 0.470 |
| Race<sup>1</sup> |  |  |  |  |
| Caucasian | 375 (97.66) | 210 (96.77) | 165 (98.80) |  |
| African American | 6 (1.56) | 4 (1.84) | 2 (1.20) | 0.272 |
| Smoking History<sup>2</sup> |  |  |  |  |
| No | 128 (33.33) | 81 (37.33) | 47 (28.14) |  |
| Yes | 251 (65.36) | 134 (61.75) | 117 (70.06) | 0.032 |
| Pack years<sup>3</sup> |  |  |  |  |
| Median (IQR) | 30 (15-45) | 30 (15-48) | 30 (13-42) | 0.926 |
| Stage (AJCC 8<sup>th</sup> Edition)<sup>4,5</sup> |  |  |  |  |
| I | 262 (68.23) | 136 (62.67) | 126 (75.45) |  |
| II | 69 (17.97) | 46 (21.20) | 23 (13.77) |  |
| III | 48 (9.90) | 21 (9.68) | 17 (10.18) |  |
| IV | 2 (0.52) | 2 (0.92) | 0 (0.00) | 0.009 |
| Treatment<sup>6</sup> |  |  |  |  |
| Surgery + Radiation + Chemo | 126 (32.81) | 74 (34.10) | 52 (31.14) |  |
| Radiation + Chemo | 107 (27.86) | 91 (41.94) | 16 (9.58) |  |
| Surgery Only | 55 (14.32) | 32 (14.75) | 23 (13.77) |  |
| Surgery + Radiation | 62 (16.15) | 14 (6.45) | 48 (28.74) |  |
| Other<sup>7</sup> | 32 (8.33) | 5 (2.30) | 27 (16.17) | <0.001 |
| HPV16 Sublineage |  |  |  |  |
| A1 | 215 (55.99) | 114 (52.53) | 101 (6.48) |  |
| A2 | 107 (27.86) | 64 (29.49) | 43 (25.75) |  |
| A3 | 3 (0.78) | 3 (1.38) | 0 (0.0) |  |
| A4 | 22 (5.73) | 14 (6.45) | 8 (4.79) |  |
| B1 | 1 (0.26) | 1 (0.46) | 0 (0.0) |  |
| C1 | 6 (1.56) | 4 (1.84) | 2 (1.20) |  |
| D1 | 1 (0.26) | 1 (0.46) | 0 (0.0) |  |
| D2 | 2 (0.52) | 0 (0.0) | 2 (1.20) |  |
| D3 | 25 (6.51) | 14 (6.45) | 11 (6.59) |  |
| D4 | 2 (0.52) | 2 (0.92) | 0 (0.0) | 0.332 |

<sup>1</sup>Race missing for 3 VUMC patients
<sup>2</sup>Smoking history missing for 5 patients; 2 from VUMC and 3 from UPMC
<sup>3</sup>Packyears missing from 73 patients; 34 from VUMC and 39 from UPMC
<sup>4</sup>Staged according to p16 result, those with missing p16 results were staged as if p16-positive
<sup>5</sup>Staging information missing for 13 patients, 12 from VUMC and 1 from UPMC
<sup>6</sup>Treatment was missing from 2 patients, 1 from VUMC and 1 from UPMC
<sup>7</sup>Other treatment includes radiation only, chemo only, surgery and chemo, palliative care, immunotherapy, clinical trial protocols
Abbreviations: VUMC, Vanderbilt University Medical Center; UPMC, University of Pittsburgh Medical Center; IQR, interquartile range; AJCC, American Joint Commission on Cancer

### Association of HPV16 SNPs with Patient Prognosis

We identified 284 site-specific variable positions (SNPs) across the 384 HPV16 genomes with a minor allele frequency (MAF) of 1.0% or greater and evaluated the association between each of these 284 HPV16 SNP and overall survival (Supplemental Table 1). Among the 384 patients included in our genomic analyses, median follow-up time was 4.00 years (interquartile range [IQR]: 2.55 to 6.78 years); 70 patients died (18.23%; median time from diagnosis to death: 2.67 years, [IQR]: 1.11 to 6.61 years). Among 284 HPV16 SNPs evaluated, 8 SNPs, varying in prevalence from 1.0% to 5.5%, were significantly associated with reduced overall survival after FDR correction (Figure 1). These 8 SNPs were localized to 4 regions within the HPV16 genome; E1 gene (position 1053); L2 gene (positions 4410, 4539, 5050, 5254), L1 gene (position 5962, 6025) and the URR (position 7173). The URR SNP was most strongly associated with an increased hazard of death (HR: 9.90 [95% CI: 3.05 to 32.12], *P*_fdr_=0.0007) (Table 2, Figure 2). Of the 384 patients, 58 (15.10%) patients had at least 1 of the 8 high-risk HPV16 SNPs; 46 patients had only 1 SNP, 10 patients had 2 SNPs, 1 patient had 3 SNPs and 1 had 4 SNPs. Patients with at least 1 of the 8 high-risk HPV16 SNPs identified had a median survival time of 3.96 years compared to 18.67 years for patients without a high-risk HPV16 SNP (log-rank test *P*<0.001); HR =7.77 (95% CI: 4.41 to 13.69, *P*=1.34×10^−12^) (Figure 3).

**Figure 1:**
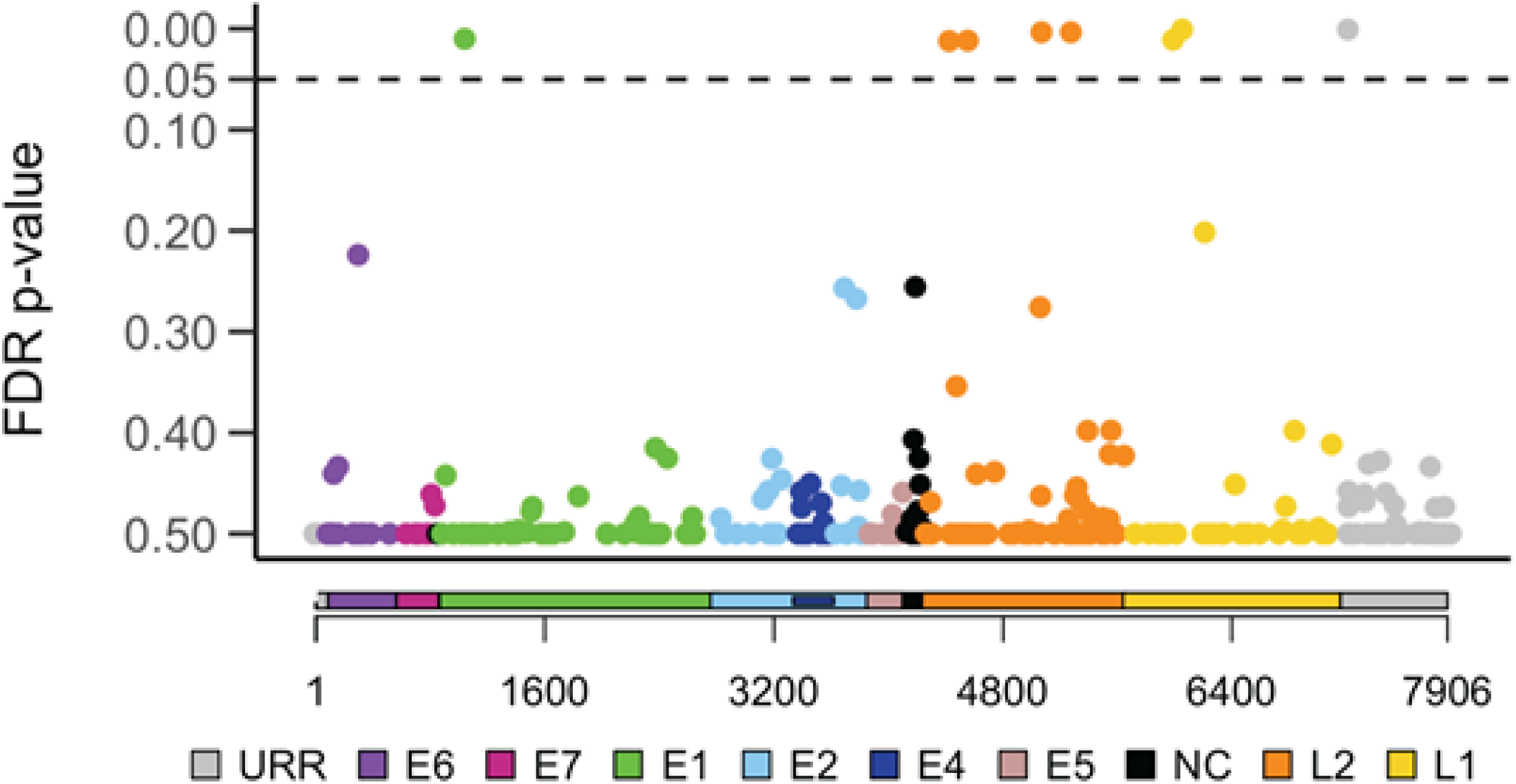
Individual HPV16 SNP associations with overall survival Cox proportional hazards models were used to generate hazard ratios and 95% confidence intervals for the risk of death (overall survival) for each minor HPV16 SNP. The major allele at each SNP position was used as the referent group. A total of 284 SNPs varied between the 388 HPV16 genomes sequenced and were included in the analysis. The Y-axis denotes the false discovery rate (FDR) corrected *P*s for the hazard ratio of each outcome. The X-axis denotes the position of each SNP in the HPV16 genome; each gene is identified with a different color. URR, upstream regulatory region (grey); E6, early gene 6 (purple); E7, early gene 7 (magenta); E1, early gene 1 (green); E2, early gene 2 (light blue); E4, early gene 4 (dark blue); E5, early gene 5 (pink); NC, non-coding region (black); L2, late gene 2 (orange); L1, late gene 1 (yellow). Dotted line denotes the FDR cutoff of 0.05.

**Table 2:** The association of HPV16 SNPs with patient prognosis

| Position | Viral Gene/Region | Minor Allele(s) | MAF (%) | Overall Survival<sup>4</sup> |  |
| --- | --- | --- | --- | --- | --- |
|  |  |  |  | Hazard Ratio (95% CI)<sup>2</sup> | *p*<sup>3</sup> |
| 1053 | E1 | C | 5.5 | 3.75 (1.77-7.95) | 0.0099 |
| 4410<sup>1</sup> | L2 | G | 2.1 | 5.32 (1.91-14.81) | 0.0120 |
| 4539 | L2 | C | 1.3 | 6.54 (2.03-21.08) | 0.0117 |
| 5050 | L2 | A/T | 2.0 | 6.53 (2.34-18.24) | 0.0030 |
| 5254 | L2 | A | 1.0 | 7.76 (2.41-24.98) | 0.0030 |
| 5962 | L1 | C | 3.4 | 4.40 (1.88-10.31) | 0.0110 |
| 6025<sup>1</sup> | L1 | T/C | 2.9 | 5.71 (2.43-13.41) | 0.0008 |
| 7173 | URR | A | 1.0 | 9.90 (3.05-32.12) | 0.0007 |

<sup>1</sup>D2 sublineage defining SNP
<sup>2</sup>Univariable Cox proportional hazards model
<sup>3</sup>False discovery rate (FDR) corrected
<sup>4</sup>384 participants were included in the analysis; 70 deaths; median follow-up time was 4.06 years (IQR: 2.25 to 6.79)
Abbreviations: MAF, minor allele frequency; CI, confidence interval

**Figure 2:**
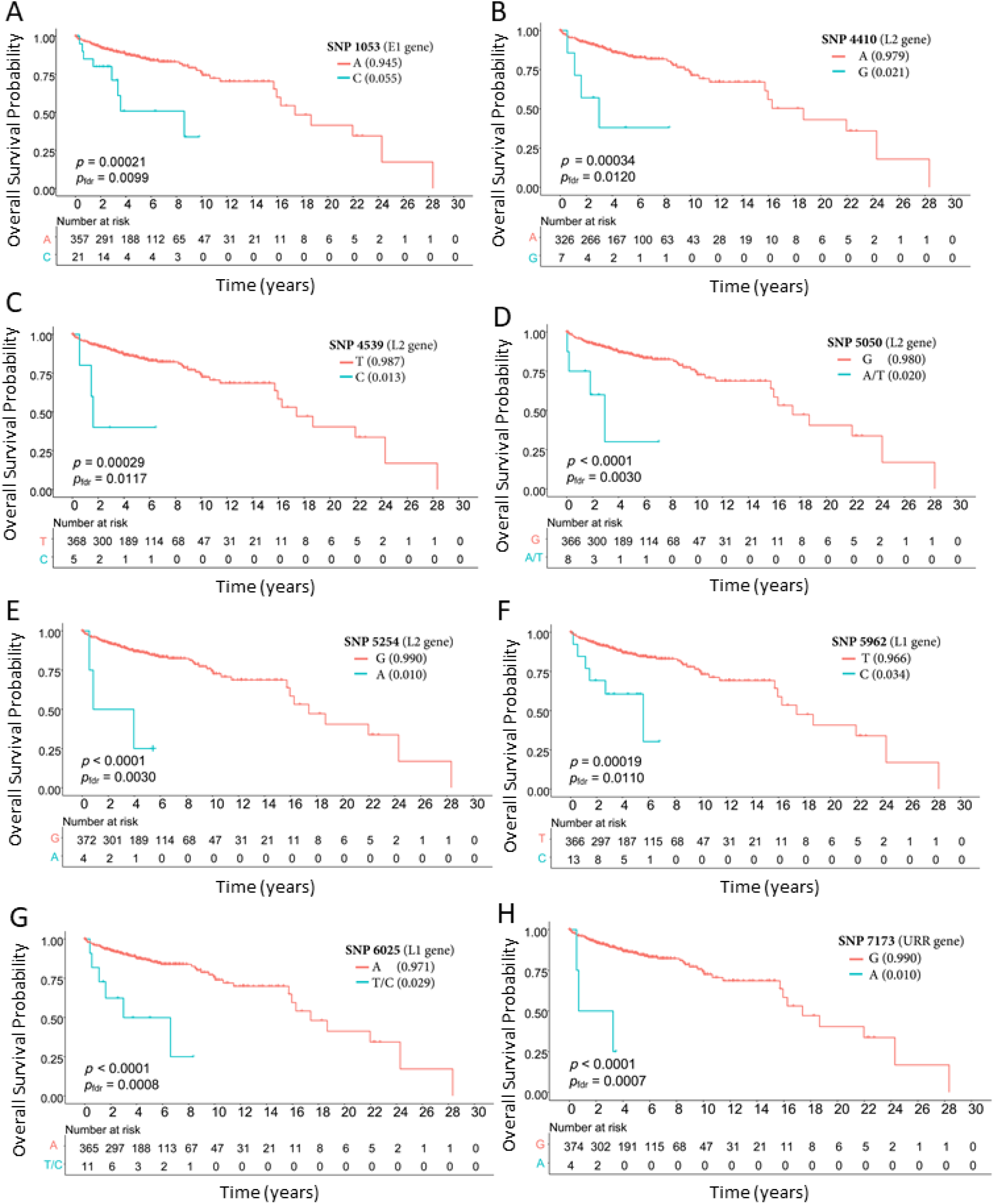
Kaplan Meier plots for overall survival by individuals high-risk HPV16 SNPs Overall survival probability comparing the minor allele (blue) to the major allele (red) for the following SNP positions within the HPV16 genome: A) 1053; B) 4410; C) 4539; D) 5050; E) 5254; E) 5962; G) 6025 and H) 7173.

**Figure 3.**
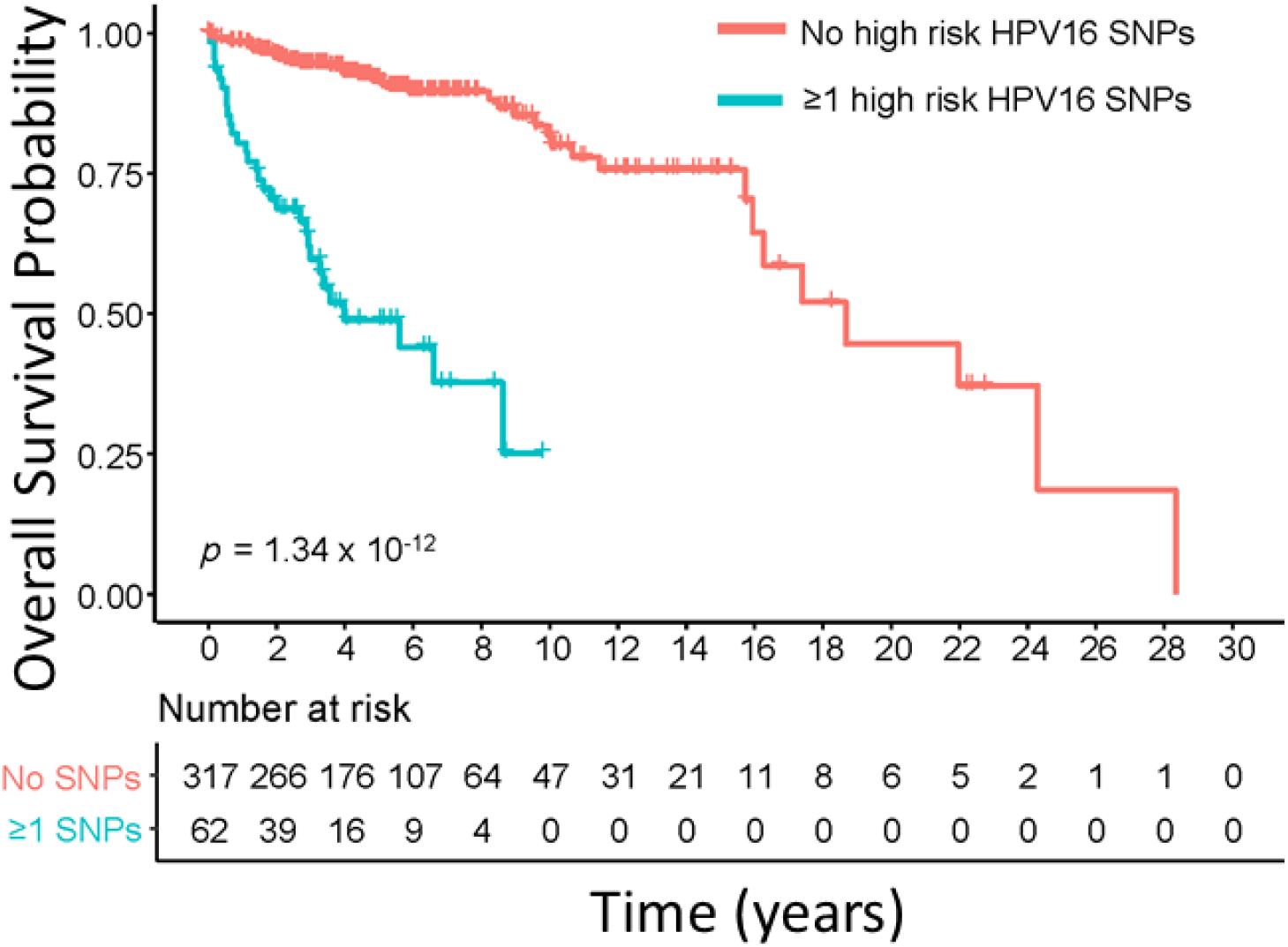
Kaplan Meier plots for overall survival comparing patients with and without at least 1 high-risk HPV16 SNP Overall survival probability comparing patients with at least 1 high-risk HPV16 SNP (blue) to patients with no high-risk HPV16 SNPs (red).

Interestingly, 2 of 8 SNPs most strongly associated with prognosis were part of the haplotype that defines the D2 sublineage (positions 4410, 6025). Eleven patients had 1 or both SNPs at positions 4410 and 6025; including both patients with the D2 sublineage and 9 patients with an A1 sublineage HPV16 virus. Of the 9 patients with an A1 sublineage, 5 had SNPs at both positions 4410 and 6025 and 4 had 6025 only. Over the course of follow-up, both D2 sublineage patients and 4 of the 9 A1 sublineage (44%) patients died compared to 18% for all 384 patients. HRs for overall survival were 5.32 (95% CI: 1.91 to 14.81, *P*_fdr_=0.0120) and 5.71 (95% CI: 2.43 to 13.41, *P*_fdr_=0.0008) for positions 4410 and 6025, respectively.

### Added Predictive Accuracy of HPV16 SNPs for Overall Survival

To evaluate the added predictive accuracy of the identified 6 high-risk SNPs, we compared patients with at least 1 of the 8 high-risk HPV16 SNPs (15.10%) to patients without a high-risk HPV16 SNP (84.90%). Compared to the Cox model with traditional factors (age, smoking, stage, treatment) alone, the Cox model that incorporated HPV16 SNP information had a significant increase in C-index of 0.069 (95% CI: 0.019 to 0.119, *P* <0.001). There were also significant increases in areas under the PPV curves for predicting 3-year (ΔAUC =0.042 [95%CI: 0.002 to 0.081], *P* =0.028), 5-year (ΔAUC =0.068 [95%CI: 0.015 to 0.111], *P* =0.008) and 8-year (ΔAUC=0.09 [95%CI: 0.026 to 0.135], *P* <0.001) overall survival, Figure 4.

**Figure 4.**
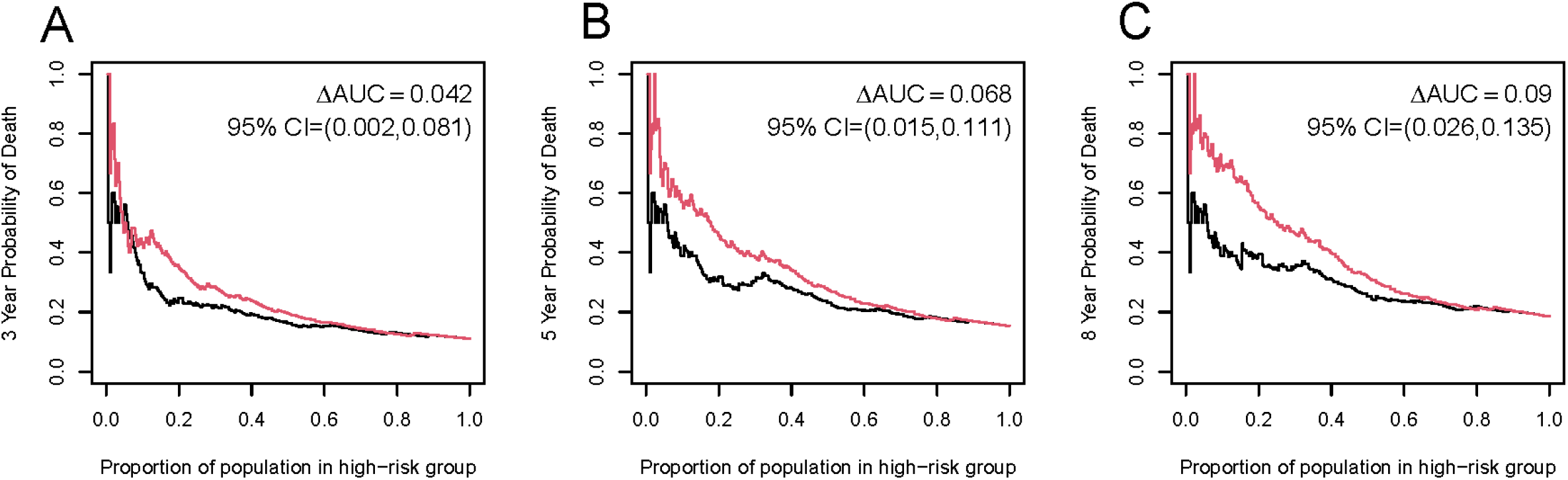
Positive Predictive Value (PPV) Curves Time-dependent PPV curves comparing the predictive accuracy of the Cox model using traditional factors (age, stage, smoking, treatment) to predict survival compared (black) to the Cox model with the addition of high-risk HPV16 SNPs (red). Panel A depicts PPV curve for predicting 3-year overall survival; Panel B for predicting 5-year overall survival and Panel C for predicting 8-year overall survival.

The HPV16 SNPs particularly improved the identification of the high-risk subgroup, i.e. patients with higher probability of death. For example, as shown in Figure 4C, patients who were categorized in the top 25% of risk using traditional factors alone had an 8-year probability of death of 34%. In contrast, patients who were categorized in the top 25% of risk using HPV16 SNPs and traditional risk factors combined had an 8-year probability of death of 54%, resulting in a 20% (95% CI: 3.1% to 37%, *P* =0.03) higher probability of death.

## DISCUSSION

To our knowledge, this is the first HPV whole genome sequencing study of HPV-OPC and patient prognosis. We used next-generation HPV16 WGS to comprehensively characterize the genetic variation within the HPV16 genome among patients with HPV-OPC. An analysis of the 284 SNPs that varied across the 384 HPV16 genomes revealed that 8 HPV16 SNPs were strongly associated with reduced overall survival. Patients with 1 or more of the 8 high-risk viral SNPs (9.8%) identified had a median survival time of only 3.96 years compared to 18.67 years for patients without a high-risk HPV16 SNP. Additionally, the inclusion of HPV16 SNP information significantly improved the predictive accuracy for HPV-OPC overall survival above traditional factors including age, smoking status, stage and treatment. This study provides the first evidence that genetics of individual HPV16 viruses may impact prognosis among patients with HPV16-driven OPC and may be a useful tool for improving risk stratification of HPV-OPC patients prior to treatment.

The 8 high-risk SNPs were located within 4 regions of the HPV16 genome: E1 gene (position 1053); L2 gene (positions 4410, 4539, 5050, 5254), L1 gene (position 5962, 6025) and the URR (position 7173). Interestingly, position 6025 is located within the FG loop of the L1 protein within the HPV16 genome and constitutes an immunodominant epitope region.^30^ Mutations within the FG surface loop of HPV16 L1 have been shown to affect recognition of both type-specific neutralizing and cross-reactive antibodies.^31, 32^ Both positions 6025 and 4410 are also considered HPV16 D2 sublineage defining variants. Prior studies in cervical cancer have consistently reported that non-European variants (A4, C1, D2, D3) were associated with an increased risk of cervical precancer and cancer, compared to the more common European sublineages (A1, A2).^33-52 17, 50, 52^ A large case-control study of 3,215 US women conducted by our group found that women infected with an HPV16 D2 sublineage had a 28 greater odds of invasive cervical cancer. Compared to the A1/A2 sublineages, the D2 sublineage was associated with a 29 greater odds of adenocarcinoma in situ and 137 greater odds of developing adenocarcinoma.^50^ Thus, prior studies of cervical cancer as well as the findings presented in this current study, suggest that SNPs more common to the D2 sublineage may confer both a greater risk of cancer development as well as contribute to more aggressive disease.

Of the 8 high-risk viral SNPs identified, position 7173 was most strongly associated with patient prognosis. Variation in the URR region near position 7173 has been previously reported in several studies of cervical cancer.^30, 53-55^ This region of the URR is postulated to contain multiple binding sites for C/EBPβ^30^ and FOXA1.^55^ Both C/EBPβ and FOXA1 are transcription factors with important roles in tumorigenesis,^55-61^ and have been shown to regulate HPV16 transcription.^62- 64^ Four of the 8 high-risk SNPs were located within the L2 gene. In addition to its structure importance, the highly conserved L2 protein is also critical for facilitating viral entry, viral DNA encapsulation, and mediating endosomal escape of the HPV genome during infection.^65^ L2 has also been implicated in facilitating immune escape by interfering with the maturation of some types of antigen presenting cells.^66^ In a large study of over 5,500 cervical specimens conducted by our group, SNPs within the URR and L2 gene regions were strongly associated with CIN3+ risk.^17^ Although the exact SNPs varied between this and our previous study, the consistency of these findings provide strong evidence for the importance of these viral regions in the carcinogenic process.

While the prevalence of each individuals high-risk HPV16 SNP ranged from 1.0% to 5.5%, combined, these SNPs were detected in 15.10% of our patient population. Having at least 1 high-risk HPV16 SNP was strongly associated with overall survival. Median survival time for patients with at least 1 high-risk HPV16 SNPs was only 3.96 years compared to 18.67 years for patients without a high-risk HPV16 SNP. Additionally, incorporating HPV16 SNP information into the Cox model resulted in significant increases in C-index and areas under the PPV curves for predicting 3-year, 5-year and 8-year overall survival compared to the Cox model that included traditional factors alone (age, stage, smoking, treatment). The HPV16 SNPs also improved the identification of individuals at high risk for poor prognosis. Taken together, the high combined prevalence of these 8 high-risk HPV16 SNPs as well as their increased predictive accuracy suggest that HPV16 SNP information may have potential clinical utility for improving risk stratification of HPV-OPC patients prior to treatment.

Our study has several strengths and limitations. We did not validate our findings in an independent cohort. Yet, it should be noted that OPC, while increasing in incidence, is rare.^9^ With a sample size of 460, this study is one of the largest studies of OPC conducted to date. Given the rarity of OPC, low rates of progression and the dominance of particular HPV16 sublineages, future studies aimed at validating our findings will likely require a consortium effort. Additionally, we focused on HPV16 given that type 16 accounts for approximately 90% of HPV-OPCs.^10^ While not the focus of the current study, samples from our study population that were HPV16-negative but high-risk HPV-positive are undergoing additional whole-genome sequencing to understand the role of other high-risk HPV types in HPV-OPC. In addition to its size, a significant strength of our study was our access to high quality clinical information with patient outcome data – this information has often been missing or incomplete in other large molecular studies of head and neck cancer.^16^ Our study was also conducted using tumors from 2 large US-based academic medical centers with robust OPC patient populations. The demographic and clinical characteristics of our patient populations closely align with those observed nationally and thus, our findings are likely to be generalizable to the larger US population.

In conclusion, this is the first study, at any anatomic site, to evaluate the association and accuracy of HPV16 genomic variation for predicting patient survival. Our findings provide the first evidence that viral variation within the HPV16 genome is associated with HPV-OPC patient prognosis and, as such, may be a potential tool for risk stratification. Additional larger studies are needed to validate these findings.

## Data Availability

All data produced in the present study are available upon reasonable request to the authors.

## Acknowledgements

This work was supported by the National Cancer Institute (NCI) K07CA218247 (PI: Krystle Kuhs) and NCI intramural funds (PI: Lisa Mirabello); Vanderbilt Clinical Oncology Research Career Development Program (K12 CA090625); National Institute for Dental and Craniofacial Research (NIDCR) K23 DE028010 (PI: Derek K. Smith) and R01 DE026471 (PI: Xiaowei Wang); Vanderbilt Ingram Cancer Center Early Detection and Prevention Program pilot grant, and the Vanderbilt Institute for Clinical and Translational Research (UL1 TR000445 from NCATS/NIH).

## Supplemental Materials

**Supplemental Table 1:** SNPs with an MAF greater than 1% and the associations with HPV-OPC overall survival (Separate file attached)

## Supplemental Figure Descriptions

**Supplemental Figure 1:**
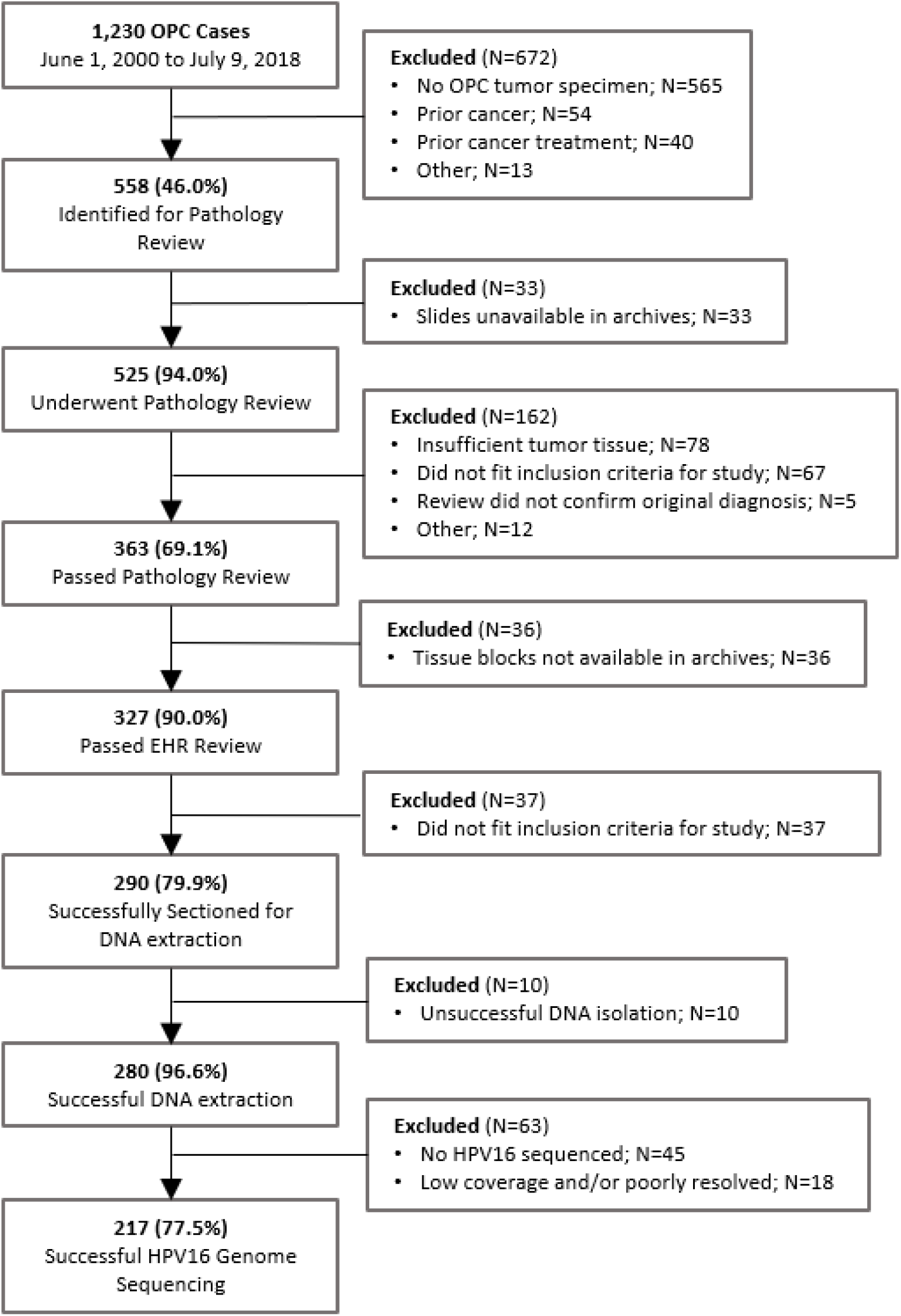
Specimen identification for VUMC.

## REFERENCES

1. Hocking JS, Stein A, Conway EL, Regan D, Grulich A, Law M, Brotherton JM. Head and neck cancer in Australia between 1982 and 2005 show increasing incidence of potentially HPV-associated oropharyngeal cancers. British journal of cancer. 2011;104(5):886–91. Epub 2011/02/03. doi: 10.1038/sj.bjc.6606091. PubMed PMID: 21285981; PMCID: 3048203.

2. Blomberg M, Nielsen A, Munk C, Kjaer SK. Trends in head and neck cancer incidence in Denmark, 1978-2007: focus on human papillomavirus associated sites. International journal of cancer Journal international du cancer. 2011;129(3):733–41. Epub 2010/09/30. doi: 10.1002/ijc.25699. PubMed PMID: 20878955.

3. Reddy VM, Cundall-Curry D, Bridger MW. Trends in the incidence rates of tonsil and base of tongue cancer in England, 1985-2006. Annals of the Royal College of Surgeons of England. 2010;92(8):655–9. Epub 2010/07/10. doi: 10.1308/003588410X12699663904871. PubMed PMID: 20615309; PMCID: 3229372.

4. Syrjanen S. HPV infections and tonsillar carcinoma. Journal of clinical pathology. 2004;57(5):449–55. Epub 2004/04/29. PubMed PMID: 15113849; PMCID: 1770289.

5. Ioka A, Tsukuma H, Ajiki W, Oshima A. Trends in head and neck cancer incidence in Japan during 1965-1999. Japanese journal of clinical oncology. 2005;35(1):45–7. Epub 2005/02/01. doi: 10.1093/jjco/hyi004. PubMed PMID: 15681605.

6. Braakhuis BJ, Visser O, Leemans CR. Oral and oropharyngeal cancer in The Netherlands between 1989 and 2006: Increasing incidence, but not in young adults. Oral oncology. 2009;45(9):e85–9. Epub 2009/05/22. doi: 10.1016/j.oraloncology.2009.03.010. PubMed PMID: 19457708.

7. Chaturvedi AK, Engels EA, Anderson WF, Gillison ML. Incidence trends for human papillomavirus-related and - unrelated oral squamous cell carcinomas in the United States. Journal of clinical oncology : official journal of the American Society of Clinical Oncology. 2008;26(4):612–9. Epub 2008/02/01. doi: 10.1200/JCO.2007.14.1713. PubMed PMID: 18235120.

8. Gillison ML, Alemany L, Snijders PJ, Chaturvedi A, Steinberg BM, Schwartz S, Castellsague X. Human papillomavirus and diseases of the upper airway: head and neck cancer and respiratory papillomatosis. Vaccine. 2012;30 Suppl 5:F34–54. Epub 2012/12/05. doi: 10.1016/j.vaccine.2012.05.070. PubMed PMID: 23199965.

9. Chaturvedi AK, Engels EA, Pfeiffer RM, Hernandez BY, Xiao W, Kim E, Jiang B, Goodman MT, Sibug-Saber M, Cozen W, Liu L, Lynch CF, Wentzensen N, Jordan RC, Altekruse S, Anderson WF, Rosenberg PS, Gillison ML. Human papillomavirus and rising oropharyngeal cancer incidence in the United States. Journal of clinical oncology : official journal of the American Society of Clinical Oncology. 2011;29(32):4294–301. Epub 2011/10/05. doi: 10.1200/JCO.2011.36.4596. PubMed PMID: 21969503; PMCID: 3221528.

10. Ndiaye C, Mena M, Alemany L, Arbyn M, Castellsague X, Laporte L, Bosch FX, de Sanjose S, Trottier H. HPV DNA, E6/E7 mRNA, and p16INK4a detection in head and neck cancers: a systematic review and meta-analysis. Lancet Oncol. 2014;15(12):1319–31. Epub 2014/12/03. doi: 10.1016/s1470-2045(14)70471-1. PubMed PMID: 25439690.

11. Jordan RC, Lingen MW, Perez-Ordonez B, He X, Pickard R, Koluder M, Jiang B, Wakely P, Xiao W, Gillison ML. Validation of methods for oropharyngeal cancer HPV status determination in US cooperative group trials. Am J Surg Pathol. 2012;36(7):945–54. Epub 2012/06/30. doi: 10.1097/PAS.0b013e318253a2d1. PubMed PMID: 22743284; PMCID: PMC6362985.

12. Ang KK, Harris J, Wheeler R, Weber R, Rosenthal DI, Nguyen-Tan PF, Westra WH, Chung CH, Jordan RC, Lu C, Kim H, Axelrod R, Silverman CC, Redmond KP, Gillison ML. Human papillomavirus and survival of patients with oropharyngeal cancer. N Engl J Med. 2010;363(1):24–35. doi: 10.1056/NEJMoa0912217. PubMed PMID: 20530316; PMCID: PMC2943767.

13. Nguyen NP, Sallah S, Karlsson U, Antoine JE. Combined chemotherapy and radiation therapy for head and neck malignancies: quality of life issues. Cancer. 2002;94(4):1131–41. PubMed PMID: 11920484.

14. Fakhry C, Zhang Q, Nguyen-Tan PF, Rosenthal D, El-Naggar A, Garden AS, Soulieres D, Trotti A, Avizonis V, Ridge JA, Harris J, Le QT, Gillison M. Human papillomavirus and overall survival after progression of oropharyngeal squamous cell carcinoma. Journal of clinical oncology : official journal of the American Society of Clinical Oncology. 2014;32(30):3365–73. doi: 10.1200/JCO.2014.55.1937. PubMed PMID: 24958820; PMCID: PMC4195851.

15. Guo T, Qualliotine JR, Ha PK, Califano JA, Kim Y, Saunders JR, Blanco RG, D’Souza G, Zhang Z, Chung CH, Kiess A, Gourin CG, Koch W, Richmon JD, Agrawal N, Eisele DW, Fakhry C. Surgical salvage improves overall survival for patients with HPV-positive and HPV-negative recurrent locoregional and distant metastatic oropharyngeal cancer. Cancer. 2015;121(12):1977–84. doi: 10.1002/cncr.29323. PubMed PMID: 25782027; PMCID: PMC4457566.

16. Cancer Genome Atlas N. Comprehensive genomic characterization of head and neck squamous cell carcinomas. Nature. 2015;517(7536):576–82. doi: 10.1038/nature14129. PubMed PMID: 25631445; PMCID: PMC4311405.

17. Mirabello L, Yeager M, Yu K, Clifford GM, Xiao Y, Zhu B, Cullen M, Boland JF, Wentzensen N, Nelson CW, Raine-Bennett T, Chen Z, Bass S, Song L, Yang Q, Steinberg M, Burdett L, Dean M, Roberson D, Mitchell J, Lorey T, Franceschi S, Castle PE, Walker J, Zuna R, Kreimer AR, Beachler DC, Hildesheim A, Gonzalez P, Porras C, Burk RD, Schiffman M. HPV16 E7 Genetic Conservation Is Critical to Carcinogenesis. Cell. 2017;170(6):1164–74 e6. Epub 2017/09/09. doi: 10.1016/j.cell.2017.08.001. PubMed PMID: 28886384; PMCID: PMC5674785.

18. Faden DL, Kuhs KAL, Lin M, Langenbucher A, Pinheiro M, Yeager M, Cullen M, Boland JF, Steinberg M, Bass S, Lewis JS, Lawrence MS, Ferris RL, Mirabello L. APOBEC Mutagenesis Is Concordant between Tumor and Viral Genomes in HPV-Positive Head and Neck Squamous Cell Carcinoma. Viruses. 2021;13(8). Epub 2021/08/29. doi: 10.3390/v13081666. PubMed PMID: 34452530; PMCID: PMC8402723.

19. Lewis JS, Jr., Mirabello L, Liu P, Wang X, Dupont WD, Plummer WD, Pinheiro M, Yeager M, Boland JF, Cullen M, Steinberg M, Bass S, Mehrad M, O’Boyle C, Lin M, Faden DL, Lang-Kuhs KA. Oropharyngeal Squamous Cell Carcinoma Morphology and Subtypes by Human Papillomavirus Type and by 16 Lineages and Sublineages. Head Neck Pathol. 2021. Epub 2021/04/03. doi: 10.1007/s12105-021-01318-4. PubMed PMID: 33797697.

20. Faden DL, Langenbucher A, Kuhs K, Lewis JS, Mirabello L, Yeager M, Boland JF, Bass S, Steinberg M, Cullen M, Lawrence MS, Ferris RL. HPV+ oropharyngeal squamous cell carcinomas from patients with two tumors display synchrony of viral genomes yet discordant mutational profiles and signatures. Carcinogenesis. 2021;42(1):14–20. Epub 2020/10/20. doi: 10.1093/carcin/bgaa111. PubMed PMID: 33075810; PMCID: PMC8014522.

21. Danciu I, Cowan JD, Basford M, Wang X, Saip A, Osgood S, Shirey-Rice J, Kirby J, Harris PA. Secondary use of clinical data: the Vanderbilt approach. J Biomed Inform. 2014;52:28–35. Epub 2014/02/19. doi: 10.1016/j.jbi.2014.02.003. PubMed PMID: 24534443; PMCID: PMC4133331.

22. Harris PA, Taylor R, Thielke R, Payne J, Gonzalez N, Conde JG. Research electronic data capture (REDCap)--a metadata-driven methodology and workflow process for providing translational research informatics support. J Biomed Inform. 2009;42(2):377–81. Epub 2008/10/22. doi: 10.1016/j.jbi.2008.08.010. PubMed PMID: 18929686; PMCID: PMC2700030.

23. Lewis JS, Jr., Beadle B, Bishop JA, Chernock RD, Colasacco C, Lacchetti C, Moncur JT, Rocco JW, Schwartz MR, Seethala RR, Thomas NE, Westra WH, Faquin WC. Human Papillomavirus Testing in Head and Neck Carcinomas: Guideline From the College of American Pathologists. Arch Pathol Lab Med. 2018;142(5):559–97. doi: 10.5858/arpa.2017-0286-CP. PubMed PMID: 29251996.

24. Cullen M, Boland JF, Schiffman M, Zhang X, Wentzensen N, Yang Q, Chen Z, Yu K, Mitchell J, Roberson D, Bass S, Burdette L, Machado M, Ravichandran S, Luke B, Machiela MJ, Andersen M, Osentoski M, Laptewicz M, Wacholder S, Feldman A, Raine-Bennett T, Lorey T, Castle PE, Yeager M, Burk RD, Mirabello L. Deep sequencing of HPV16 genomes: A new high-throughput tool for exploring the carcinogenicity and natural history of HPV16 infection. Papillomavirus Research. 2015;1:3–11. doi: http://dx.doi.org/10.1016/j.pvr.2015.05.004.

25. Cingolani P, Platts A, Wang LL, Coon M, Nguyen T, Wang L, Land SJ, Lu X, Ruden DM. A program for annotating and predicting the effects of single nucleotide polymorphisms, SnpEff: SNPs in the genome of Drosophila melanogaster strain w(1118); iso-2; iso-3. Fly. 2012;6(2):80–92. doi: 10.4161/fly.19695. PubMed PMID: PMC3679285.

26. Stamatakis A. RAxML-VI-HPC: maximum likelihood-based phylogenetic analyses with thousands of taxa and mixed models. Bioinformatics. 2006;22(21):2688–90. Epub 2006/08/25. doi: 10.1093/bioinformatics/btl446. PubMed PMID: 16928733.

27. Harrell FE, Jr., Lee KL, Califf RM, Pryor DB, Rosati RA. Regression modelling strategies for improved prognostic prediction. Stat Med. 1984;3(2):143–52. Epub 1984/04/01. doi: 10.1002/sim.4780030207. PubMed PMID: 6463451.

28. Zheng Y, Cai T, Pepe MS, Levy WC. Time-dependent Predictive Values of Prognostic Biomarkers with Failure Time Outcome. J Am Stat Assoc. 2008;103(481):362–8. Epub 2008/01/01. doi: 10.1198/016214507000001481. PubMed PMID: 19655041; PMCID: PMC2719907.

29. Chen L, Lin DY, Zeng D. Predictive accuracy of covariates for event times. Biometrika. 2012;99(3):615–30. Epub 2013/07/12. doi: 10.1093/biomet/ass018. PubMed PMID: 23843671; PMCID: PMC3635702.

30. Gurgel AP, Chagas BS, do Amaral CM, Nascimento KC, Leal LR, Silva Neto Jda C, Cartaxo Muniz MT, de Freitas AC. Prevalence of human papillomavirus variants and genetic diversity in the L1 gene and long control region of HPV16, HPV31, and HPV58 found in North-East Brazil. Biomed Res Int. 2015;2015:130828. Epub 2015/03/21. doi: 10.1155/2015/130828. PubMed PMID: 25793187; PMCID: PMC4352477.

31. Carpentier GS, Fleury MJ, Touze A, Sadeyen JR, Tourne S, Sizaret PY, Coursaget P. Mutations on the FG surface loop of human papillomavirus type 16 major capsid protein affect recognition by both type-specific neutralizing antibodies and cross-reactive antibodies. J Med Virol. 2005;77(4):558–65. Epub 2005/10/29. doi: 10.1002/jmv.20492. PubMed PMID: 16254978.

32. Bissett SL, Godi A, Beddows S. The DE and FG loops of the HPV major capsid protein contribute to the epitopes of vaccine-induced cross-neutralising antibodies. Sci Rep. 2016;6:39730. Epub 2016/12/23. doi: 10.1038/srep39730. PubMed PMID: 28004837; PMCID: PMC5177933.

33. Burk RD, Harari A, Chen Z. Human papillomavirus genome variants. Virology. 2013;445(1–2):232–43. doi: http://dx.doi.org/10.1016/j.virol.2013.07.018.

34. Hildesheim A, Schiffman M, Bromley C, Wacholder S, Herrero R, Rodriguez AC, Bratti MC, Sherman ME, Scarpidis U, Lin Q-Q, Terai M, Bromley RL, Buetow K, Apple RJ, Burk RD. Human Papillomavirus Type 16 Variants and Risk of Cervical Cancer. Journal of the National Cancer Institute. 2001;93(4):315–8. doi: 10.1093/jnci/93.4.315.

35. Pientong C, Wongwarissara P, Ekalaksananan T, Swangphon P, Kleebkaow P, Kongyingyoes B, Siriaunkgul S, Tungsinmunkong K, Suthipintawong C. Association of human papillomavirus type 16 long control region mutation and cervical cancer. Virology Journal. 2013;10(1):30. PubMed PMID: doi:10.1186/1743-422X-10-30.

36. Xi LF, Koutsky LA, Hildesheim A, Galloway DA, Wheeler CM, Winer RL, Ho J, Kiviat NB. Risk for High-Grade Cervical Intraepithelial Neoplasia Associated with Variants of Human Papillomavirus Types 16 and 18. Cancer Epidemiology Biomarkers & Prevention. 2007;16(1):4–10. doi: 10.1158/1055-9965.epi-06-0670.

37. Schiffman M, Rodriguez AC, Chen Z, Wacholder S, Herrero R, Hildesheim A, Desalle R, Befano B, Yu K, Safaeian M, Sherman ME, Morales J, Guillen D, Alfaro M, Hutchinson M, Solomon D, Castle PE, Burk RD. A Population-Based Prospective Study of Carcinogenic Human Papillomavirus Variant Lineages, Viral Persistence, and Cervical Neoplasia. Cancer Research. 2010;70(8):3159–69. doi: 10.1158/0008-5472.can-09-4179.

38. Cornet I, Gheit T, Iannacone MR, Vignat J, Sylla BS, Del Mistro A, Franceschi S, Tommasino M, Clifford GM. HPV16 genetic variation and the development of cervical cancer worldwide. British journal of cancer. 2013;108(1):240–4. Epub 2012/11/22. doi: 10.1038/bjc.2012.508. PubMed PMID: 23169278; PMCID: PMC3553516.

39. Gheit T, Cornet I, Clifford GM, Iftner T, Munk C, Tommasino M, Kjaer SK. Risks for Persistence and Progression by Human Papillomavirus Type 16 Variant Lineages Among a Population-Based Sample of Danish Women. Cancer Epidemiology Biomarkers & Prevention. 2011;20(7):1315–21. doi: 10.1158/1055-9965.epi-10-1187.

40. Zehbe I, Voglino G, Delius H, Wilander E, Tommasino M. Risk of cervical cancer and geographical variations of human papillomavirus 16 E6 polymorphisms. The Lancet. 1998;352(9138):1441–2. doi: 10.1016/S0140-6736(05)61263-9.

41. Zuna RE, Moore WE, Shanesmith RP, Dunn ST, Wang SS, Schiffman M, Blakey GL, Teel T. Association of HPV16 E6 variants with diagnostic severity in cervical cytology samples of 354 women in a US population. International Journal of Cancer. 2009;125(11):2609–13. doi: 10.1002/ijc.24706.

42. Sichero L, Ferreira S, Trottier H, Duarte-Franco E, Ferenczy A, Franco EL, Villa LL. High grade cervical lesions are caused preferentially by non-European variants of HPVs 16 and 18. International Journal of Cancer. 2007;120(8):1763–8. doi: 10.1002/ijc.22481.

43. Berumen J, Ordoñez RM, Lazcano E, Salmeron J, Galvan SC, Estrada RA, Yunes E, Garcia-Carranca A, Gonzalez-Lira G, Madrigal-de la Campa A. Asian-American Variants of Human Papillomavirus 16 and Risk for Cervical Cancer: a Case–Control Study. Journal of the National Cancer Institute. 2001;93(17):1325–30. doi: 10.1093/jnci/93.17.1325.

44. Freitas LB, Chen Z, Muqui EF, Boldrini NAT, Miranda AE, Spano LC, Burk RD. Human Papillomavirus 16 Non-European Variants Are Preferentially Associated with High-Grade Cervical Lesions. PLoS ONE. 2014;9(7):e100746. doi: 10.1371/journal.pone.0100746. PubMed PMID: PMC4077691.

45. Burk RD, Terai M, Gravitt PE, Brinton LA, Kurman RJ, Barnes WA, Greenberg MD, Hadjimichael OC, Fu L, McGowan L, Mortel R, Schwartz PE, Hildesheim A. Distribution of Human Papillomavirus Types 16 and 18 Variants in Squamous Cell Carcinomas and Adenocarcinomas of the Cervix. Cancer Research. 2003;63(21):7215–20.

46. Quint KD, de Koning MNC, van Doorn L-J, Quint WGV, Pirog EC. HPV genotyping and HPV16 variant analysis in glandular and squamous neoplastic lesions of the uterine cervix. Gynecologic Oncology. 2010;117(2):297–301. doi: 10.1016/j.ygyno.2010.02.003. PubMed PMID: 20207397.

47. Rabelo-Santos SH, Villa LL, Derchain SF, Ferreira S, Sarian LOZ, Ângelo-Andrade LAL, Do Amaral Westin MC, Zeferino LC. Variants of human papillomavirus types 16 and 18: Histological findings in women referred for atypical glandular cells or adenocarcinoma in situ in cervical smear. International Journal of Gynecological Pathology. 2006;25(4):393–7. doi: 10.1097/01.pgp.0000215302.17029.0c.

48. De Boer MA, Peters LAW, Aziz MF, Siregar B, Cornain S, Vrede MA, Jordanova ES, Fleuren GJ. Human papillomavirus type 18 variants: Histopathology and E6/E7 polymorphisms in three countries. International Journal of Cancer. 2005;114(3):422–5. doi: 10.1002/ijc.20727.

49. Lizano M, De la Cruz-Hernández E, Carrillo-García A, García-Carrancá A, Ponce de Leon-Rosales S, Dueñas-González A, Hernández-Hernández DM, Mohar A. Distribution of HPV16 and 18 intratypic variants in normal cytology, intraepithelial lesions, and cervical cancer in a Mexican population. Gynecologic Oncology. 2006;102(2):230–5. doi: http://dx.doi.org/10.1016/j.ygyno.2005.12.002.

50. Mirabello L, Yeager M, Cullen M, Boland JF, Chen Z, Wentzensen N, Zhang X, Yu K, Yang Q, Mitchell J, Roberson D, Bass S, Xiao Y, Burdett L, Raine-Bennett T, Lorey T, Castle PE, Burk RD, Schiffman M. HPV16 Sublineage Associations With Histology-Specific Cancer Risk Using HPV Whole-Genome Sequences in 3200 Women. J Natl Cancer Inst. 2016;108(9). doi: 10.1093/jnci/djw100. PubMed PMID: 27130930.

51. Hirose Y, Onuki M, Tenjimbayashi Y, Yamaguchi-Naka M, Mori S, Tasaka N, Satoh T, Morisada T, Iwata T, Kiyono T, Mimura T, Sekizawa A, Matsumoto K, Kukimoto I. Whole-Genome Analysis of Human Papillomavirus Type 16 Prevalent in Japanese Women with or without Cervical Lesions. Viruses. 2019;11(4). Epub 2019/04/19. doi: 10.3390/v11040350. PubMed PMID: 30995759; PMCID: PMC6520816.

52. Clifford GM, Tenet V, Georges D, Alemany L, Pavon MA, Chen Z, Yeager M, Cullen M, Boland JF, Bass S, Steinberg M, Raine-Bennett T, Lorey T, Wentzensen N, Walker J, Zuna R, Schiffman M, Mirabello L. Human papillomavirus 16 sub-lineage dispersal and cervical cancer risk worldwide: Whole viral genome sequences from 7116 HPV16-positive women. Papillomavirus Res. 2019;7:67–74. Epub 2019/02/10. doi: 10.1016/j.pvr.2019.02.001. PubMed PMID: 30738204; PMCID: PMC6374642.

53. Dai S, Li C, Yan Z, Zhou Z, Wang X, Wang J, Sun L, Shi L, Yao Y. Association of Human Papillomavirus Type 16 Long Control Region Variations with Cervical Cancer in a Han Chinese Population. Int J Med Sci. 2020;17(7):931–8. Epub 2020/04/21. doi: 10.7150/ijms.43030. PubMed PMID: 32308546; PMCID: PMC7163361.

54. Martinelli M, Villa C, Sotgiu G, Muresu N, Perdoni F, Musumeci R, Combi R, Cossu A, Piana A, Cocuzza C. Analysis of Human Papillomavirus (HPV) 16 Variants Associated with Cervical Infection in Italian Women. Int J Environ Res Public Health. 2020;17(1). Epub 2020/01/08. doi: 10.3390/ijerph17010306. PubMed PMID: 31906371; PMCID: PMC6982298.

55. Xi J, Chen J, Xu M, Yang H, Luo J, Pan Y, Wang X, Qiu L, Yang J, Sun Q. Genetic variability and functional implication of the long control region in HPV-16 variants in Southwest China. PLoS One. 2017;12(8):e0182388. Epub 2017/08/03. doi: 10.1371/journal.pone.0182388. PubMed PMID: 28767682; PMCID: PMC5540483.

56. Tsukada J, Yoshida Y, Kominato Y, Auron PE. The CCAAT/enhancer (C/EBP) family of basic-leucine zipper (bZIP) transcription factors is a multifaceted highly-regulated system for gene regulation. Cytokine. 2011;54(1):6–19. Epub 2011/01/25. doi: 10.1016/j.cyto.2010.12.019. PubMed PMID: 21257317.

57. Cowper-Sal lari R, Zhang X, Wright JB, Bailey SD, Cole MD, Eeckhoute J, Moore JH, Lupien M. Breast cancer risk-associated SNPs modulate the affinity of chromatin for FOXA1 and alter gene expression. Nat Genet. 2012;44(11):1191–8. Epub 2012/09/25. doi: 10.1038/ng.2416. PubMed PMID: 23001124; PMCID: PMC3483423.

58. Li Z, Tuteja G, Schug J, Kaestner KH. Foxa1 and Foxa2 are essential for sexual dimorphism in liver cancer. Cell. 2012;148(1-2):72–83. Epub 2012/01/24. doi: 10.1016/j.cell.2011.11.026. PubMed PMID: 22265403; PMCID: PMC3266536.

59. Jin HJ, Zhao JC, Ogden I, Bergan RC, Yu J. Androgen receptor-independent function of FoxA1 in prostate cancer metastasis. Cancer Res. 2013;73(12):3725–36. Epub 2013/03/30. doi: 10.1158/0008-5472.CAN-12-3468. PubMed PMID: 23539448; PMCID: PMC3686855.

60. Lin L, Miller CT, Contreras JI, Prescott MS, Dagenais SL, Wu R, Yee J, Orringer MB, Misek DE, Hanash SM, Glover TW, Beer DG. The hepatocyte nuclear factor 3 alpha gene, HNF3alpha (FOXA1), on chromosome band 14q13 is amplified and overexpressed in esophageal and lung adenocarcinomas. Cancer Res. 2002;62(18):5273–9. Epub 2002/09/18. PubMed PMID: 12234996.

61. Wang J, Bao W, Qiu M, Liao Y, Che Q, Yang T, He X, Qiu H, Wan X. Forkhead-box A1 suppresses the progression of endometrial cancer via crosstalk with estrogen receptor alpha. Oncol Rep. 2014;31(3):1225–34. Epub 2014/01/24. doi: 10.3892/or.2014.2982. PubMed PMID: 24452315.

62. Struyk L, van der Meijden E, Minnaar R, Fontaine V, Meijer I, ter Schegget J. Transcriptional regulation of human papillomavirus type 16 LCR by different C/EBPbeta isoforms. Mol Carcinog. 2000;28(1):42–50. Epub 2000/05/23. PubMed PMID: 10820487.

63. Bernard HU. Regulatory elements in the viral genome. Virology. 2013;445(1-2):197–204. Epub 2013/06/04. doi: 10.1016/j.virol.2013.04.035. PubMed PMID: 23725692.

64. Sichero L, Sobrinho JS, Villa LL. Identification of novel cellular transcription factors that regulate early promoters of human papillomavirus types 18 and 16. J Infect Dis. 2012;206(6):867–74. Epub 2012/06/29. doi: 10.1093/infdis/jis430. PubMed PMID: 22740717.

65. Doorbar J, Egawa N, Griffin H, Kranjec C, Murakami I. Human papillomavirus molecular biology and disease association. Rev Med Virol. 2015;25 Suppl 1:2–23. Epub 2015/03/11. doi: 10.1002/rmv.1822. PubMed PMID: 25752814; PMCID: PMC5024016.

66. Fahey LM, Raff AB, Da Silva DM, Kast WM. A major role for the minor capsid protein of human papillomavirus type 16 in immune escape. J Immunol. 2009;183(10):6151–6. Epub 2009/10/30. doi: 10.4049/jimmunol.0902145. PubMed PMID: 19864613; PMCID: PMC2947488.

